# Reduction in initiations of HIV treatment in South Africa during the COVID pandemic

**DOI:** 10.1101/2021.08.18.21262046

**Authors:** Mariet Benade, Lawrence Long, Sydney Rosen, Gesine Meyer-Rath, Jeanne-Marie Tucker, Jacqui Miot

## Abstract

**Background:** In response to the global pandemic of COVID-19, countries around the world began imposing stay-at-home orders, restrictions on transport, and closures of businesses in early 2020. South Africa implemented a strict lockdown in March 2020 before its first COVID-19 wave started, gradually lifted restrictions between May and September 2020, and then re-imposed restrictions in December 2020 in response to its second wave. There is concern that COVID-19-related morbidity and mortality, fear of transmission, and government responses may have led to a reduction in antiretroviral treatment (ART) initiations for HIV-infected individuals in countries like South Africa.

**Methods:** We analyzed national, public sector, facility-level data from South Africa’s District Health Information System (DHIS) from January 2019 to March 2021 to quantify changes in ART initiation rates stratified by province, setting, facility size and type and compared the timing of these changes to COVID-19 case numbers and government lockdown levels. We excluded facilities with missing data, mobile clinics, and correctional facilities. We estimated the total number of ART initiations per study month for each stratum and compared monthly totals, by year.

**Results:** At the 2471 facilities in the final data set (59% of all ART sites in the DHIS), 28% fewer initiations occurred in 2020 than in 2019. Numbers of ART initiations declined sharply in all provinces in April-June 2020, compared to the same months in 2019, and remained low for the rest of 2020, with some recovery between COVID-19 waves in October 2020 and possible improvement beginning in March 2021. Percentage reductions were largest in district hospitals, larger facilities, and urban areas. After the initial decline in April-June 2020, most provinces experienced a clear inverse relationship between COVID-19 cases and ART initiations but little relationship between ART initiations and lockdown level.

**Conclusions:** The COVID-19 pandemic and responses to it resulted in substantial declines in the number of HIV-infected individuals starting treatment in South Africa, with no recovery of numbers during 2020. These delays may lead to worse treatment outcomes for those with HIV and potentially higher HIV transmission. Exceptional effort will be needed to sustain gains in combating HIV.

## Background

The global pandemic of COVID-19 has disrupted many aspects of life for people around the world, including access to care for chronic diseases such as HIV. Alarms were raised early on about how COVID-19-related illness, fear of COVID-19 transmission, and national and local measures to prevent transmission, such as “lockdowns,” would harm timely access to HIV treatment [1]. While medications continued to be provided to many experienced antiretroviral therapy (ART) patients through home or community delivery[2], those needing to initiate or re-initiate ART usually have to attend an established clinic or hospital. There is reason for serious concern that global targets for initiating new HIV-positive patients on treatment were not met in 2020 and may be missed in 2021[3, 4].

South Africa’s first wave of COVID-19 began in June 2020 and ended in August 2020; a second wave was recognized from December 2020 to January 2021 and a third wave began in May 2021, with provincial variations[5]. South Africa, which continues to have nearly three million HIV-positive residents not yet on ART[6], declared a national state of disaster on 15 March 2020 and implemented strict lockdown measures such as stay-at-home orders, travel restrictions, and the closure of schools, non-essential businesses, and most public transportation on 27 March. Restrictions were gradually lifted in increments between 1 May and 21 September 2020 before being tightened again in late December 2020[7]. During the initial lockdown, individuals were permitted to leave their homes to seek healthcare, but public transportation was not readily available, and many primary healthcare clinics temporarily closed or reduced operating hours due to staff infections and exposures requiring quarantine, other staff absences, stockouts of supplies, and/or redeployment of resources for COVID-19 screening and testing. [8]. Fear of COVID-19 transmission also deterred routine clinic access, and a sharp downturn in testing for HIV during the lockdowns likely reduced the number of potential ART initiation candidates[8].

Although a few articles and reports have discussed the potential impact of the COVID-19 restrictions on HIV service delivery in South Africa[9], we are aware of only one empirical study, conducted during the first half of 2020 in a single province, that estimated actual changes in initiation numbers in a large population. Among 65 public primary care clinics in urban and rural KwaZulu-Natal, a 34% decrease in ART initiations/week was observed between the average over the two years preceding the start of the lockdown and the three months following it, after which the weekly average number of initiations increased again, reaching 82.7% of pre-lockdown levels at the end of July 2020[10]. To provide a geographically broader and more detailed understanding of the impact of the COVID-19 pandemic and pandemic control measures and improve future decision making, we used national data to quantify changes in ART initiation rates between January 2019 and March 2021 by province and to identify facility characteristics associated with larger or smaller changes.

## Methods

### Data

The District Health Information System (DHIS)[11] is used by the South African Government to collect aggregate (facility-level) data about service delivery at the more than 4,000 public sector facilities in the country, which provide HIV services for roughly 92% of South Africa’s population[12]. Facilities report monthly on a wide range of process indicators, including the number of individuals initiated (or reinitiated) on ART. Reporting is generally fairly complete; although multi-month lags in data entry for individual facilities are common, backlogs are generally cleared in the subsequent months. The accuracy of the data reported has been shown to vary among sites[13].

We accessed national DHIS data for 01 January 2019 to 31 March 2021 for all facilities in the DHIS data set that reported at least one month of ART initiation data during the study period. We then excluded from the dataset facilities that did not have observations for every month of the study period (i.e. that had data missing for one or more of the 27 months studied). We also excluded mobile clinics and correctional facilities. Our final analytic data set included one observation—the number of patients initiated on ART—for each facility for each of the 27 months in the study period. We also collected information on the province, size, setting (urban, peri-urban, or rural), and type (clinic, hospital, or community health centres) for each facility[14]. Facility size was defined as either “small” (<1000 total patients retained on ART in 2019) or “large” (≥1000 ART patients in 2019) as per DHIS data on ART retention at each site.

In addition to DHIS data, we used publicly-available data on monthly COVID-19 cases in each province and on the dates at which different levels of lockdown were imposed[5].

### Analysis

We first calculated the total number of ART initiations per study month and year for the country as a whole and for each province and compared totals and percentage changes for each calendar month in each of the study years, with 2019 as the baseline. For example, the number of initiations in March 2019 was compared to that in March 2020 and March 2021, to control for seasonal differences in healthcare utilization unrelated to COVID-19. We then stratified the data set by province, district, facility type, size, and setting and compared changes in ART initiation numbers between the groups to determine if specific subsets of facilities experienced larger or smaller changes in service volumes. Finally, we examined the relationships over time among ART initiations, COVID cases, and lockdown levels.

## Results

### Study population

Of the 4,200 ART facilities included in the original data set, we excluded 1,360 that were missing data and 369 additional nontraditional treatment sites (338 mobile sites and 31 correctional facilities), leaving an analytic data set of 2471 facilities, equivalent to 59% of those in the original data set. Facilities included in the analysis were located in all 9 provinces and included a mix of sizes, settings, and facility types, as shown in Table 1.

**Table 1.** Characteristics of facilities included in the analysis (n=2,471)

| Characteristic | Total sites in DHIS data set | Included in analysis (n, % of total sites in DHIS data set) | Total number of patients retained on ART at the facilities included in 2019 (column %) | Total number of ART initiations at the facilities included in 2019 (column %) |
| --- | --- | --- | --- | --- |
| All fixed-site facilities | 4,200 | 2,471 (59) | 4,017,406 | 636,969 |
| *Facility type (public sector only)* |  |  |  |  |
| Primary healthcare clinics | 3109 | 2041 (66) | 3,008,830 (75) | 463,924 (73) |
| Community health centres | 332 | 270 (81) | 806,707 (20) | 139,028 (22) |
| Hospitals | 283 | 160 (57) | 201,868 (5) | 34,017 (5) |
| Mobile sites and correctional facilities (excluded) | 759 | 0 (0) | n.a. | n.a. |
| *Number of patients on ART\** |  |  |  |  |
| <1000 | 2734 | 1079 (39) | 640,487 (16) | 91,190 (14) |
| ≥ 1000 | 1466 | 1392 (95) | 3,376,918 (84) | 545,779 (86) |
| *Setting* |  |  |  |  |
| Urban | 1814 | 1229 (68) | 2,561,859 (64) | 435,911 (68) |
| Peri-urban | 378 | 198 (52) | 304,590 (8) | 46,581 (7) |
| Rural | 2008 | 1044 (52) | 1,150,956 (29) | 154,477 (24) |
| *Province* |  |  |  |  |
| Eastern Cape | 888 | 411 (46) | 381,516 (9) | 61,167 (10) |
| Free State | 294 | 178 (61) | 247,121 (6) | 32,060 (5) |
| Gauteng | 446 | 371 (84) | 995,382 (25) | 187,531 (29) |
| KwaZulu Natal | 840 | 547 (65) | 1,199,113 (30) | 176,714 (28) |
| Limpopo | 539 | 329 (61) | 282,360 (7) | 36,547 (6) |
| Mpumalanga | 340 | 267 (79) | 432,045 (11) | 71,490 (11) |
| Northern Cape | 183 | 50 (27) | 33,895 (1) | 3,902(1) |
| North West | 380 | 192 (51) | 230,501 (6) | 37,856 (6) |
| Western Cape | 290 | 126 (44) | 215,474 (5) | 29,702 (5) |

\*Mean number of patients retained in 2019

### Numbers of ART initiations compared to 2019

Figure 1 illustrates the total number of patients initiated on ART per month for the full study period.

**Figure 1.**
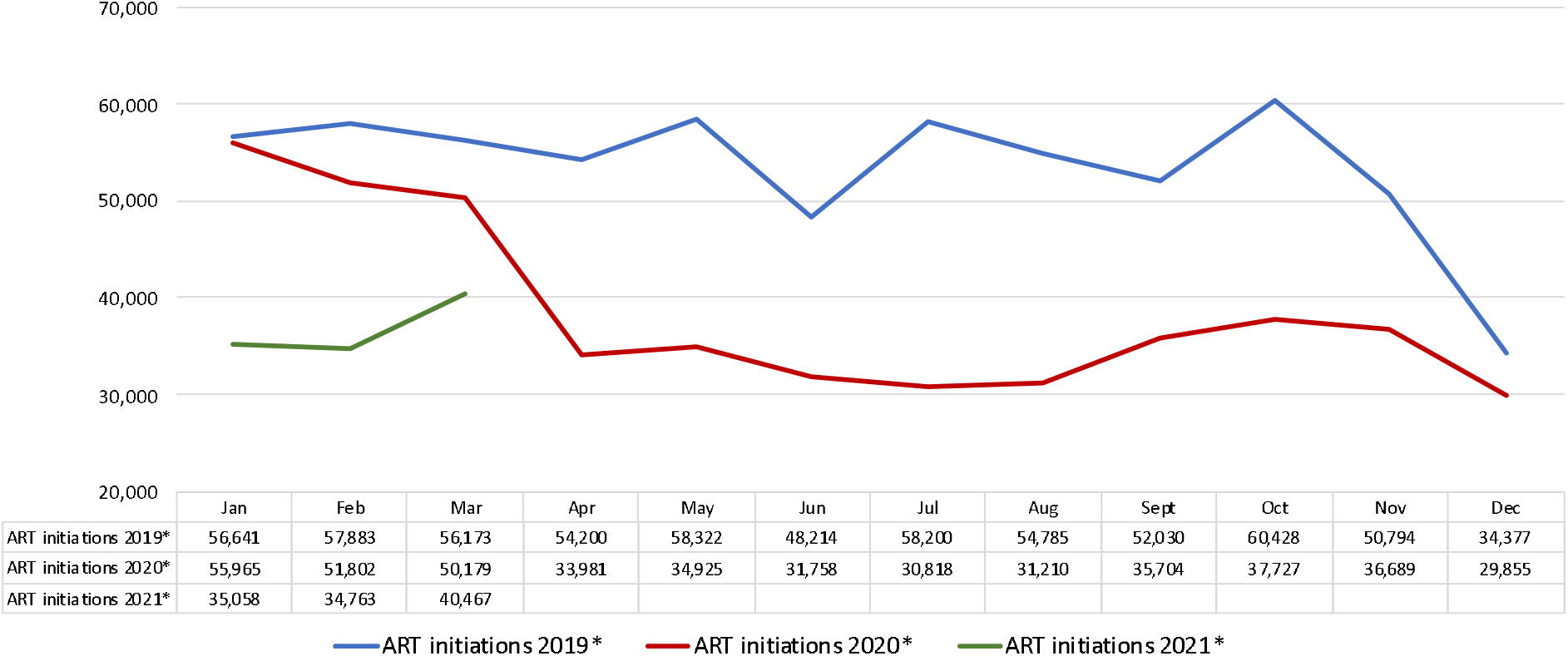
ART initiations in South Africa, January 2019-March 2021. *Includes 2,471 facilities; excludes facilities with missing data, mobile clinics, and correctional facilities

Countrywide, the number of patients initiated on ART was similar in January 2019 and 2020. It began to fall off modestly in February and March 2020. There was then a steep decline in April and May 2020, compared to the same months in 2019. This discrepancy persisted for the rest of 2020, with some recovery from September to November 2020, prior to the country’s second wave of COVID-19, and possible improvement beginning in March 2021. At the facilities included in our analysis, which comprise 59% of all ART facilities in South Africa, a total of 180,681 fewer patients initiated ART in 2020 than in 2019, or a reduction of 28%.

In Figure 2, we illustrate the number of ART initiations in calendar year 2020 and the first quarter of 2021 in each province of South Africa as percentages of the numbers of initiations in each quarter of 2019.

**Figure 2.**
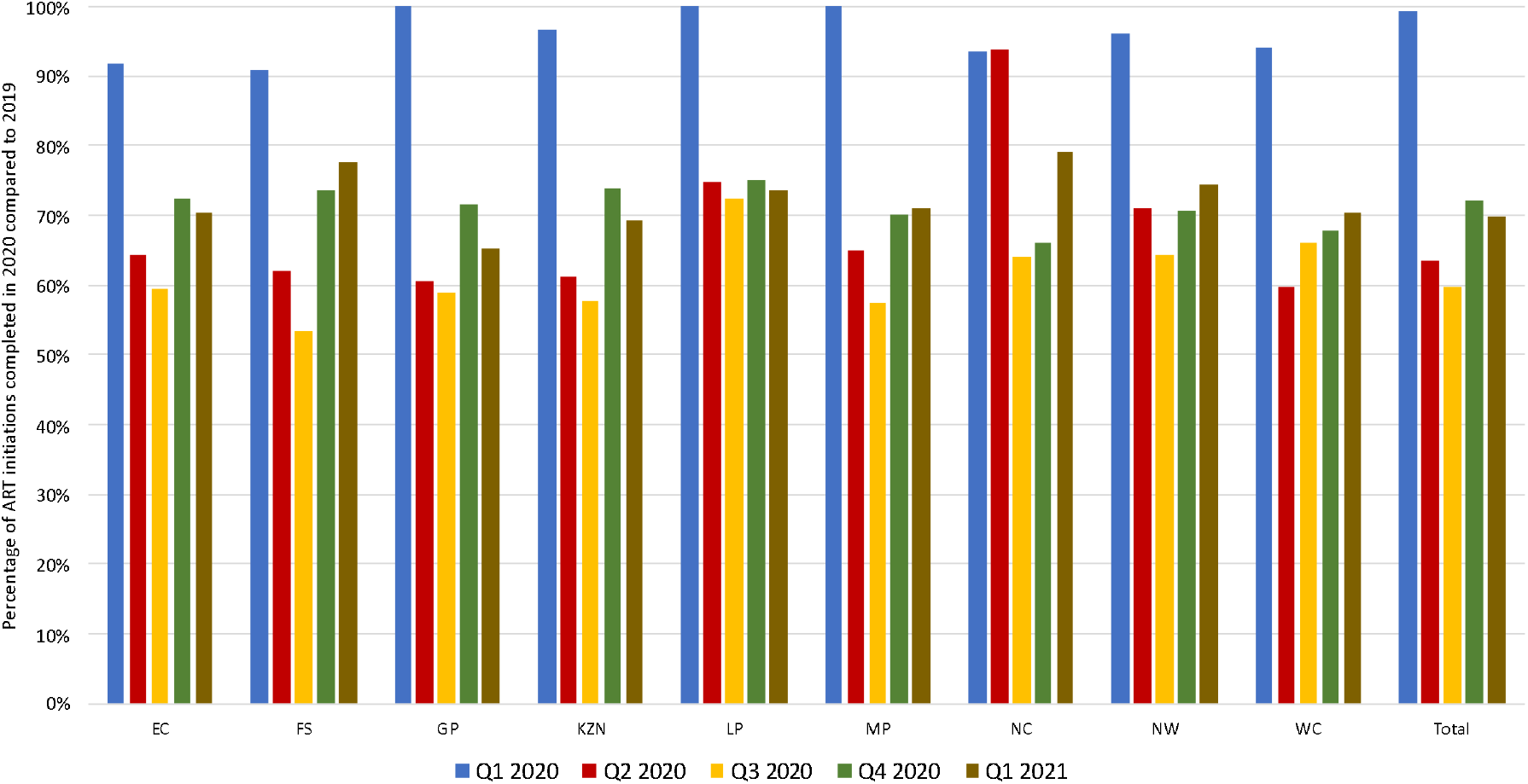
ART initiations in 2020 as a percentage of the number of initiations in 2019, by province.

All provinces experienced a sharp drop-off between the first and second quarters of 2020 seen in Figure 2. After that, while patterns were roughly consistent across the provinces, the timing and magnitude of the changes differed, with the fastest decline and earliest recovery in the Western Cape Province and a slower decline in the Eastern Cape and Free State provinces, among others.

We next looked at the changes in ART initiation numbers by key facility characteristics, as shown in Figure 3.

**Figure 3.**
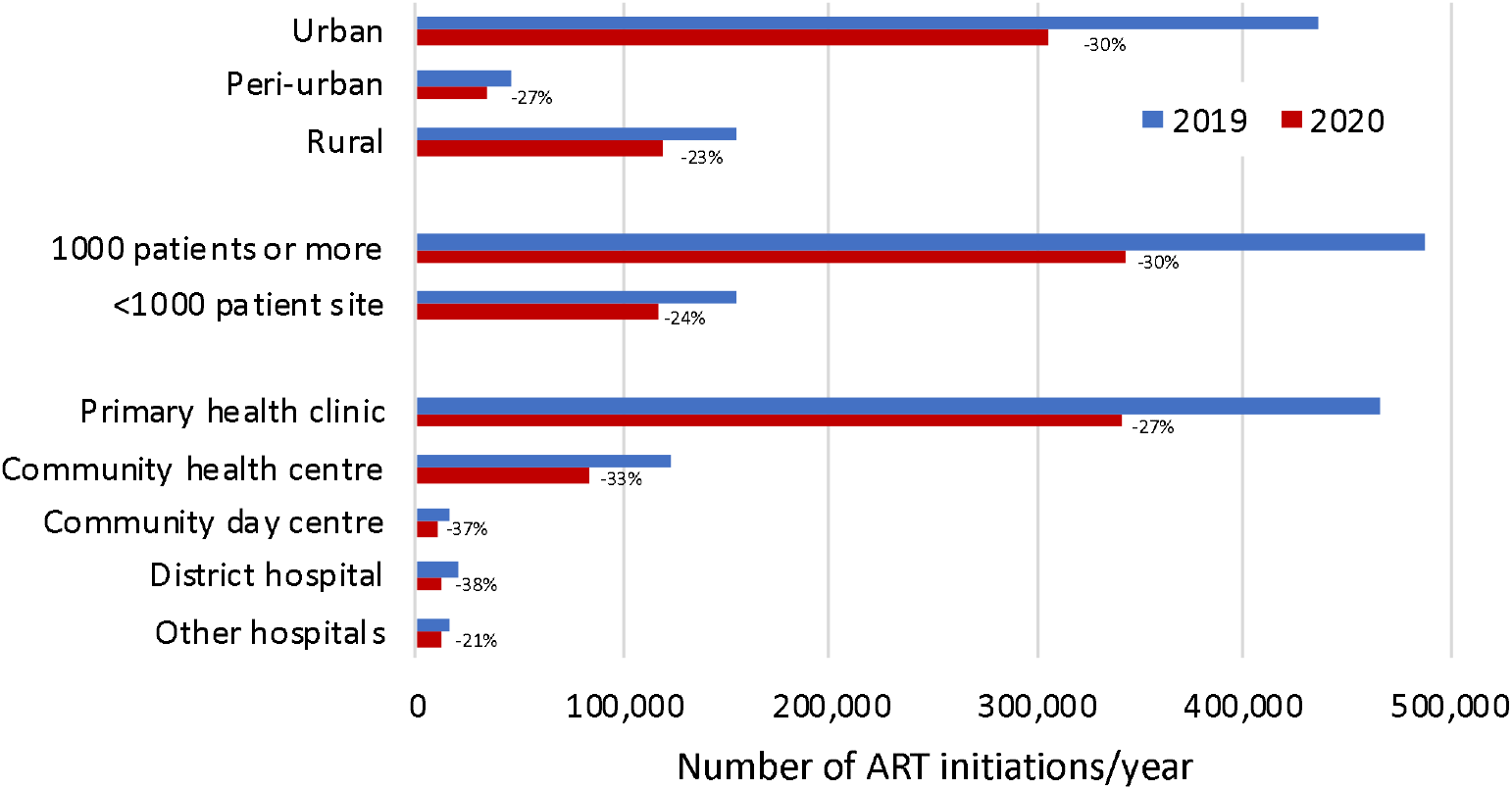
ART initiations in 2019 and 2020 by setting, size, and facility type.

Reductions in initiation numbers were largest in percentage terms in district hospitals, larger facilities, and urban areas. Primary health clinics, which are responsible for 75% of all initiations in the country, experienced a decrease of 27% in 2020.

### Changes in ART initiation numbers, COVID-19 cases, and lockdown levels

Finally, we compared the timing of ART initiations, COVID-19 cases, and national lockdown restrictions. In Figure 4, these variables are presented for the country as a whole; provincial data are shown in Figure 5.

**Figure 4.**
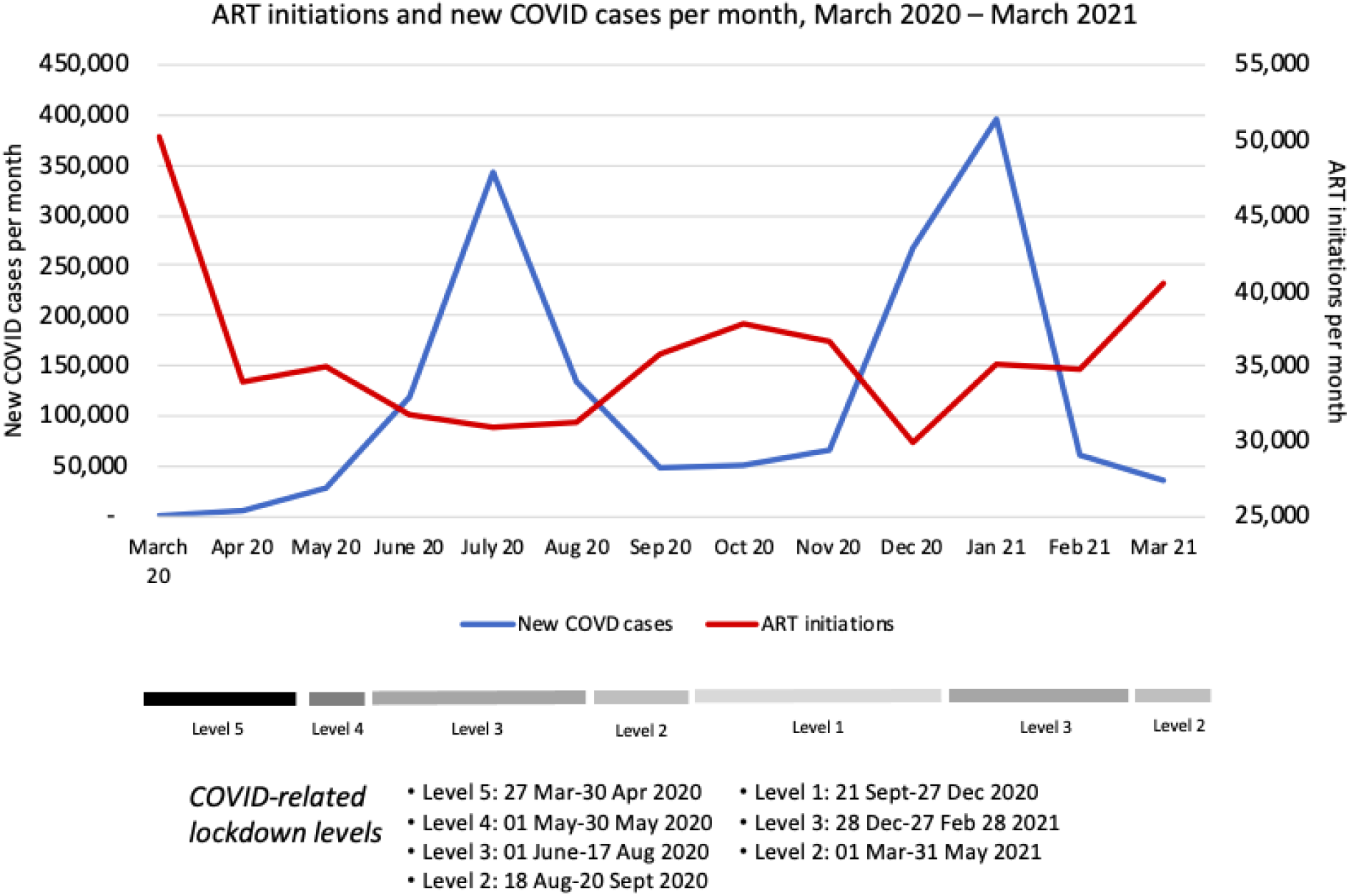
ART initiations and new COVID-19 cases per month, March 2020-March 2021.

**Figure 5.**
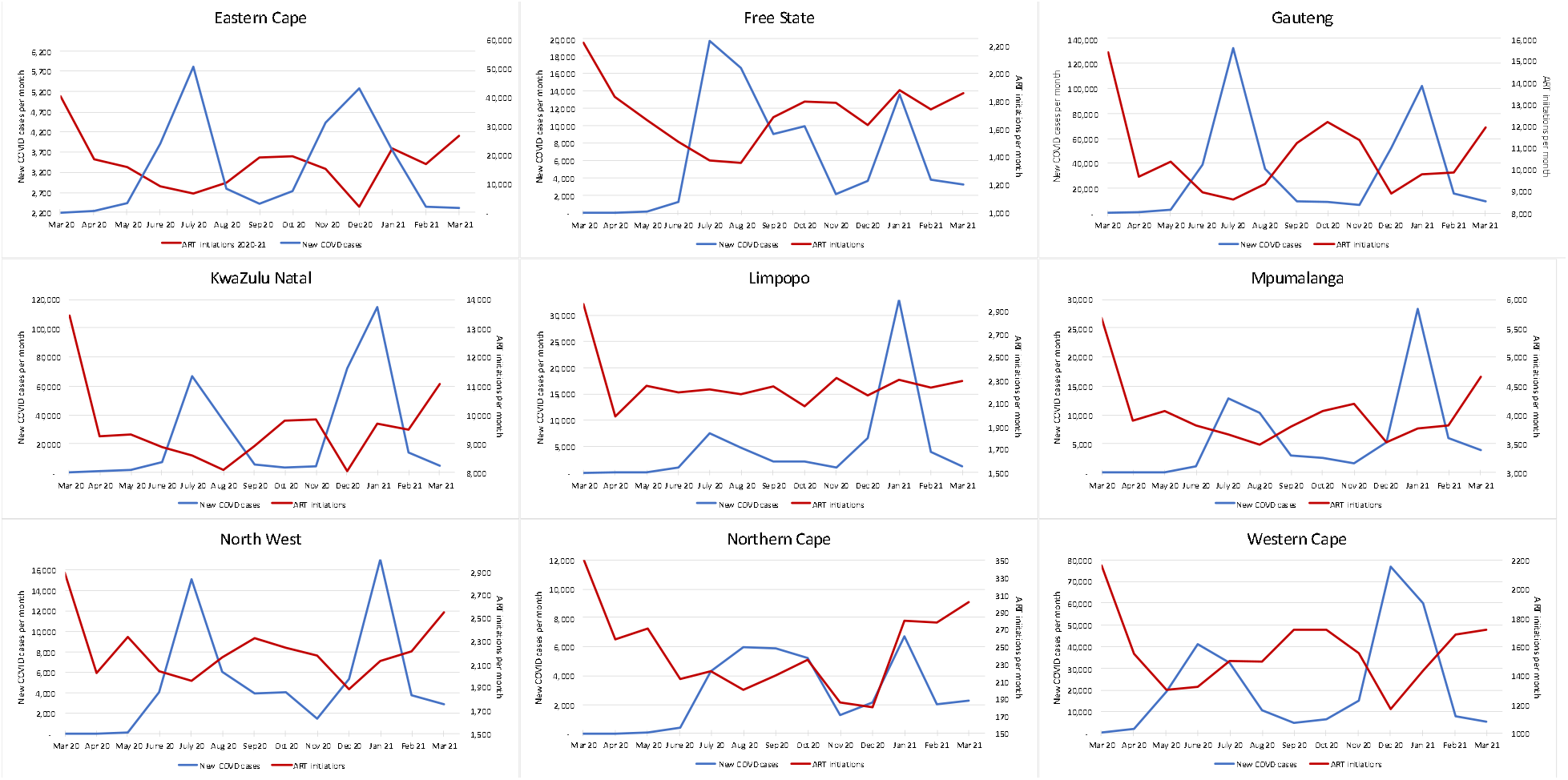
ART initiations and new COVID-19 cases per month by province, March 2020-March 2021.

A sharp decline in ART initiations was seen in every province at the time of the imposition of the level 5 lockdown in late March 2020, or even slightly before, potentially responding to the declaration of a state of disaster in mid-March. Diagnosed cases of COVID-19 began to rise steeply only in May 2020, after the shift to lockdown level 4, which eased some restrictions. After the initial decline, most provinces experienced a clear inverse relationship between COVID-19 cases and ART initiations but little relationship between ART initiations and lockdown level.

## Discussion

In this analysis of aggregate data reported by roughly three fifths of the public sector, ART-providing healthcare facilities in South Africa, we found that the number of HIV-positive individuals initiating ART fell substantially in 2020 from the number in 2019. ART initiations fell in all provinces and for all facility types, sizes, and settings. Although there was some recovery between the first and second COVID-19 waves, in October-November 2020, numbers remained well below those seen in 2019. After the initial drop-off in April-May 2020, which preceded South Africa’s first wave of COVID-19, ART initiations generally fell as diagnosed COVID-19 infections rose, rather than in direct proportion to the intensity of the national lockdown.

The data set used in this analysis covered 59% of South African public sector ART facilities. If the overall relative decline in ART initiations estimated for the study sample is extrapolated to 100% of the public sector ART facilities in the country, we calculate that some 310,000 fewer initiations took place in 2020 than in 2019, a reduction of about 28%. In its 2020/21 Annual Performance Plan, issued before the start of the COVID-19 pandemic, the National Department of Health in South Africa aimed to add a total of 1.17 million patients to its national ART program over the course of 2020[15]. If we again extrapolate our results to the entire country, we find that in 2020 the performance targets were underachieved by roughly 375,000 initiations, or 32% of the stated target.

The impact of this discrepancy on South Africa’s HIV program and HIV-related morbidity and mortality rates will depend on whether the missing patients will come forward for ART initiation in 2021 or 2022, the health system’s capacity to provide services to these patients, and the extent to which HIV transmission increases as a result of HIV-positive patients spending more time off ART. With the arrival of South Africa’s third wave of COVID-19 in June 2021 and the corresponding re-imposition of lockdowns, it seems likely that low uptake of ART will continue until at least the second half of 2021 and possibly well until 2022, depending on vaccine coverage and case rates. In this case, many of the patients who do ultimately present for ART initiation will have more advanced disease and lower CD4 counts than they otherwise would have had, potentially threatening their prognosis on ART[16], while some will have died of HIV-related causes before starting ART. These deaths, due indirectly to the COVID-19 pandemic, likely comprise some of the many non-COVID “excess deaths” reported for the country since March 2020[17]. There is also some evidence that HIV-positive individuals with low CD4 counts—including many of those who faced COVID-19-related delays in initiating ART—face a greater risk of mortality if they are infected with COVID-19[18].

As mentioned above, we found only one other published study that estimated changes in numbers of ART initiations for South Africa. In KwaZulu Natal Province, Dorward et al (2021), also using DHIS data, observed a similar pattern of a sharp downturn in April-May 2020, followed by some recovery up to June, when their data set ended. In contrast to our results, however, they found no difference between rural and urban areas of KZN Province, while we observed a larger effect in urban areas. (Of interest, we also note that Dorward et al reported that the recovery in ART initiation numbers they saw in June and July 2020 was primarily among female patients; initiations among males remained low[10]. We were not able to assess differences by sex with our data set.)

Our study had a number of limitations. First, as explained above, our analysis included only public sector health facilities in the DHIS that reported complete data (monthly observations) for the entire study period. The selection bias introduced by the exclusion of roughly 2/5 of the country’s facilities is unclear, but we speculate that it may have biased the sample in favor of better-resourced and/or better-performing sites. Second, we had no way to validate the data in the DHIS database, but as noted above, we know from others’ analyses that accuracy is variable. Third, our study period ended in March 2021, just after the end of South Africa’s second wave of COVID-19. We do not know if ART initiation numbers recovered over the course of April and May, prior to the start of the third wave in June 2021. It is possible that, given a longer study period, ART initiation may have recovered to pre-lockdown levels. Fourth, our comparison values, from 2019, may or may not be a valid counter-factual for 2020. In some areas, ART initiation rates were trending down prior to the pandemic, as the number of HIV-infected persons not on ART declined. On the other hand, efforts made to achieve the Government’s 2020/2021 goals for HIV treatment coverage may have led to a larger number of ART initiations in 2020, in the absence of the pandemic. Fifth, it is possible that some of the patients who would have initiated ART in the public sector in 2020 instead accessed private sector facilities, though we have no evidence of this happening.

Finally, our data cannot tell us whether the observed reduction in initiations reflects COVID-19 morbidity, fear of COVID-19 transmission, reduction in the health system’s capacity to provide services, or constraints on movement due to the lockdown. We believe that it was almost certainly a combination of these factors, though the timing of the trends in initiations suggests that less can be attributed to official lockdowns, and more to the pandemic itself, than may be commonly thought. Regardless of the causes, exceptional effort and investment will be needed in the coming months to protect the massive gains that have been made in protecting the health and lives of those infected and affected by HIV.

## Data Availability

Data used in this analysis are from the South African District Health Information System (DHIS II) and are owned by the South African National Department of Health. These data cannot be made publicly available by the authors. DHIS data access can be requested from the Director-General of the National Department of Health in South Africa at the following address:
Director: National Health Information System
Cluster: Health Information, Research, Monitoring & Evaluation
Civitas Building
Private Bag X 828
Pretoria, 0001, South Africa
Data on COVID-19 case numbers are available from https://mediahack.co.za/datastories/coronavirus/dashboard/.

## Acknowledgements

We are grateful to Matthew P. Fox of Boston University for his input on this analysis.

